# Implementation of a clinical decision support tool for acute diarrhea management in Tanzania and the United States: A Qualitative study using the Consolidated Framework for Implementation Research

**DOI:** 10.64898/2026.08.20.26360926

**Authors:** Joan Chepngeno, Rochelle K. Rosen, Ryan Lantini, Stephanie C. Garbern, Mwemezi Salvatory, Raban Rameck, Fatimah Dhalla, Dayoung Yu, Vikram Sharma, Christopher Duggan, Karim P. Manji, Adam C. Levine

## Abstract

**Background:** In two large studies conducted in Bangladesh, our recently developed artificial intelligence (AI)-based models for assessing dehydration severity in children under five years (DHAKA models) and patients over age five (NIRUDAK models) were significantly more accurate and reliable than the WHO IMCI and IMAI guidelines for diarrhea management. We incorporated these models into a novel mobile health (mHealth) clinical decision support tool (CDST), called “FluidCalc”, with the potential to improve acute diarrhea management by frontline health workers worldwide. Our objective was to assess the barriers and facilitators to uptake and use of our mHealth CDST in both a low-resource setting (Tanzania) and high-resource setting (United States (US)) among healthcare providers and stakeholders.

**Methods:** Qualitative data were collected through focus group discussions (FGDs) with healthcare providers and in-depth interviews (IDIs) with stakeholders and policymakers from February - July 2025 in Tanzania and February - March 2026 in the US. The Consolidated Framework for Implementation Research (CFIR) was used to guide discussions and elicit participant feedback. Audio recordings were transcribed and translated from Swahili to English where applicable, and data were analyzed using framework matrix analysis.

**Results:** 35 providers from different cadres participated in FGDs, and 13 stakeholders participated in IDIs. Facilitators to implementation included FluidCalc’s simplicity, ease of use, and offline functionality. Participants reported that the app could streamline clinical workflows, promote adherence to diarrhea management guidelines, facilitate task shifting, support antibiotic stewardship, and reduce errors in fluid rehydration calculations. FluidCalc was also viewed as a valuable teaching tool, and for supporting less experienced healthcare providers and trainees, and as useful during diarrheal disease outbreaks. Perceived barriers included the need for reliable digital infrastructure, including access to mobile devices, internet connectivity, and dependable electricity and lengthy institutional approval processes. Endorsement and approval from the Ministry of Health and health facility leadership were perceived as essential for successful implementation.

**Conclusion:** Healthcare providers and stakeholders believe FluidCalc has the potential to improve care for patients with acute diarrhea in both high- and low-resource settings. Addressing identified barriers and ensuring reliable digital health infrastructure are needed to support effective integration into patient care.

**Contributions to literature:**

- FluidCalc is a clinical decision support tool (CDST) for managing acute diarrhea.
- Few implementation studies have examined CDSTs for diarrhea management across both high-resource and low-resource settings. This study provides comparative insights into factors influencing implementation in two distinct health system contexts.
- Beyond technical performance and usefulness, CDST uptake depends on clinicians’ perceptions of the tool’s impact on workflow, workload, clinical autonomy, and compatibility with existing health systems.
- Additionally, organizational context shapes perceptions of digital innovations, helping explain why a tool may be perceived as an opportunity to improve care or as an added implementation burden.

## Background

Diarrheal diseases continue to be a leading cause of morbidity and mortality worldwide, particularly among young children and populations living in low- and middle-income countries (LMICs). In 2019, diarrheal illness accounted for 370,000 deaths among children under five and contributed substantially to global disability and health system strain [1,2]. Although mortality rates have declined over time, the burden remains disproportionately concentrated in resource-limited settings and LMICs, where constraints on safe water, sanitation and challenges on access and availability to public health infrastructure persist [2]. Importantly, diarrheal diseases are not limited to early childhood and contribute to significant morbidity, healthcare utilization, and preventable mortality across all age groups [3]. Indeed, over 381 million cases of diarrhea and 569,000 diarrheal deaths occur annually in patients over 65 years of age around the world, representing a 200% increase in cases of diarrhea in the elderly over the past 30 years [3]. Even in high resource settings like the United States (US), diarrhea accounts for 179 million outpatient visits, 500,000 hospitalizations, and 5,000 deaths each year [4].

The effective management of acute diarrhea relies heavily on timely assessment and appropriate rehydration as the foundation of treatment [5,6]. Accurate assessment of dehydration severity is crucial to guide fluid administration, prevent complications from both under- and over-hydration, and improve clinical outcomes, especially in low-resource settings where close monitoring of fluid status is challenging [7,8]. However, existing clinical decision frameworks, such as the widely used World Health Organization (WHO) Integrated Management of Childhood Illness (IMCI) and Integrated Management of Adolescent and Adult Illness (IMAI) algorithms, have shown important limitations in real-world settings. Studies have shown limited diagnostic accuracy, with high false-positive rates for moderate and severe dehydration, raising concerns about the effectiveness of continuing to use existing guidelines as the standard-of-care [9–12]. These limitations contribute to gaps in quality of care and highlight the need for more reliable and scalable tools to support frontline decision-making in the management of diarrheal illness [13,14].

In response to these challenges, digital health and mobile health (mHealth), have emerged as a promising approach to support clinical decision-making, triage, training, and communication across a wide range of healthcare settings [15,16]. Mobile clinical decision support tools (CDSTs) can improve the accuracy of provider assessments, enhance adherence to guidelines, and support standardized care delivery at the point of care [17–20]. These tools also show a promising approach for task-shifting to community health workers (CHWs) and non- physician providers, improving access to care where workforce shortages and infrastructure constraints limit traditional healthcare delivery [21–23]. As digital health technologies rapidly mature and expand globally, they represent an increasingly important strategy for strengthening health systems and improving outcomes for high-burden conditions such as diarrheal disease [15,16]. However, their effectiveness, usability, and implementation in real-world clinical contexts remain areas of active investigation [23, 24].

FluidCalc is a CDST for dehydration management in patients with acute diarrhea. The app is available for free via www.FluidCalc.org and on the Google Play Store and iOS App Store as a mobile application, incorporating the *Novel, Innovative Research for Understanding Dehydration in Adults and Kids* (NIRUDAK) and *Dehydration: Assessing Kids Accurately* (DHAKA) clinical diagnostic models for assessing dehydration severity in patients with acute diarrhea aged under five years and over five years, respectively [25]. Previous research conducted in Bangladesh showed that the DHAKA and NIRUDAK models are more accurate than the current IMCI and IMAI algorithms recommended by the World Health Organization [ 26, 27]. Qualitative research has also been conducted to test the clinical utility of the mobile application itself among frontline nurses and physicians; feedback from participants was overall positive and informed revisions to the mobile CDST, but these perspectives were also limited to clinicians in Bangladesh [25].

In order to successfully implement novel mHealth tools, it is crucial to understand healthcare workers’ perceptions regarding these tools in diverse settings, as well as engage providers in the adoption process [28]. Using an implementation science-based approach, this study aims to evaluate potential barriers and facilitators to uptake and usage of the FluidCalc CDST by a variety of health providers and stakeholders working in a low resource setting (Tanzania) and in a high resource setting (United States). By identifying context-specific factors that influence implementation, this work will inform the external validation of the FluidCalc CDST across new patient populations and clinical settings.

## Methods

### Study Population and Setting

Purposeful sampling was used to enroll health professionals and stakeholders in clinical management of acute diarrhea in Tanzania and the United States. At the Tanzania site, participants were enrolled from four health facilities in Dar es Salaam: Temeke General Hospital, Buza Health Center, Round Table Health Center and Mbagala Rangi Tatu Health Center. In the US, participants were recruited for virtual focus group discussions using a purposeful sample of health providers working in primary care, urgent care, and emergency care settings across the United States.

### Data Collection

Each site used a prepared qualitative topic guide which covered specific questions related to the CFIR constructs (see Appendix 1 and 2). The topic guide included intent statements and questions linked to the relevant CFIR domains. Links to download the app or use a browser-based version of the app were sent to participants in advance of the interview or focus group, and a live demonstration of the FluidCalc app was conducted for each focus group discussion (FGD) or in-depth interview (IDI). Facilitators sought specific feedback from participants about app use, including the utility of the treatment recommendations (intervention characteristics); advantages and disadvantages to incorporating FluidCalc into their clinical work flow and environment, including who should complete data entry into the app and who should implement rehydration recommendations (inner setting/individuals); who would determine how the app is used and what strategies would help with implementation and training (implementation process); and what existing protocols or policies would need to be followed to ensure implementation (outer setting). We also asked about use of the FluidCalc app during relevant outbreak scenarios (Cholera in Tanzania) or (norovirus in the United States).

FGDs and IDIs were conducted by local study staff trained in qualitative facilitation, the CFIR, and the study goals in Tanzania, and by the principal investigator (ACL) and co-investigators (JC, RKR, SCG, RL) in the US. Written informed consent was obtained from all participants. All FGDs and IDIs were audio recorded, transcribed and, where necessary, translated into English from Swahili. Transcriptions were reviewed against the audio files for accuracy, correct speaker attribution and any needed deidentification. Participants were identified in transcripts using a study identification number.

### Data Analysis

Transcript data was summarized using framework matrices. In this data reduction analysis protocol, a framework is created with participants represented in rows and key questions/codes in columns [29]. Members of the data analysis team reviewed the transcripts and summarized participant comments about each question/code in the appropriate cell. As a credibility step, all transcripts, and the data charted from them in the framework matrix, were reviewed by at least two team members. Corrections and additions made to the matrix by the second reviewer were discussed with the full research team. Additionally, because of the clinical content of the discussions, at least one of the reviewers was a currently practicing physician who could speak to the clinical context of app use in Tanzania or the United States.

After data was charted, we created a summary of each key topic and research question. These summaries were also reviewed and discussed by the US research team and used to develop the themes presented below.

## Results

### Characteristics of participants

A total of 48 participants were included in the study. In Tanzania, participants included general practice physicians (*n*=4), nurses (*n*=4), pharmacists (*n*=3), clinical officers (*n*=4), community health workers (*n*=5), and stakeholders and policymakers (*n*=6). In the United States, participants included emergency care physicians caring for adults (*n*=4), emergency care physicians caring for children (*n*=3), primary care physicians caring for adults (*n*=4), primary care physicians caring for children (*n*=4), and stakeholders and policymakers (*n*=7).

Full characteristics of the participants are shown in Tables 1 and 2.

**Table 1.** Description of focus group participants.

| Focus group # | Country | Age in years (range) | Gender |  | Occupation |
| --- | --- | --- | --- | --- | --- |
| | | | Female ($n=17$) | Male ($n=18$) | |
| 1 | Tanzania | 29-56 | 0 | 4 | Clinical officers |
| 2 | Tanzania | 35-44 | 2 | 2 | General practice physicians |
| 3 | Tanzania | 29-56 | 3 | 2 | Community health workers |
| 4 | Tanzania | 34-45 | 4 | 0 | Nurses |
| 5 | Tanzania | 29-51 | 1 | 2 | Pharmacists |
| 6 | United States | 32-66 | 1 | 3 | Adult emergency care physicians |
| 7 | United States | 43-59 | 2 | 1 | Pediatric emergency care physicians |
| 8 | United States | 36-47 | 2 | 2 | Adult primary care physicians |
| 9 | United States | 36-69 | 2 | 2 | Pediatric primary care physicians |

**Table 2.** Description of in-depth interview participants.

| Interview # | Country | Age in years (Range) | Gender | Occupation and years in current role |
| --- | --- | --- | --- | --- |
| 1 | Tanzania | 30-39 | Female | Medical officer in charge; 5 years in current role |
| 2 | Tanzania | 30-39 | Male | Acting district medical officer, district AIDS, STIs and Hepatitis coordinator; < 1 year in current role |
| 3 | Tanzania | 30-39 | Male | Head of emergency medicine department; 7 years in current role |
| 4 | Tanzania | 40-49 | Female | Clinical services coordinator and acting medical officer in charge; 7 years in current role |
| 5 | Tanzania | 50-59 | Male | Chief medical officer of the province; 8 years in current role |
| 6 | Tanzania | 50-59 | Male | Medical officer in charge; 5 years in current role |
| 7 | United States | 60-69 | Female | Chair of family medicine at academic medical center; 3 years in current role |
| 8 | United States | 60-69 | Male | Global health informatics researcher focused on electronic health record systems and clinical decision support, and diagnostic/triage apps; approximately 8–9 years in current role |
| 9 | United States | 40-49 | Female | Attending emergency medicine physician at a level 1 trauma center and community emergency department; approximately 9 years in current role |
| 10 | United States | 60-69 | Male | Pediatric emergency medicine physician working in the emergency department focusing on children; approximately 26 years of clinical experience |
| 11 | United States | 40-49 | Male | Infectious diseases physician and clinic director of an infectious diseases and immunology clinic; 16 years as an infectious disease physician |
| 12 | United States | 50-59 | Male | Senior WHO officer supporting ministries of health to plan models of health care delivery and providing support for clinical areas such as trauma and sepsis; 5 years in current role |
| 13 | United States | 40-49 | Female | Family nurse practitioner at a community health center urgent care; 5 years in current role |

### Prior exposure to digital health tools

Participants reported using a range of digital tools to support patient care, including both standalone smartphone-based applications and those integrated into electronic health records (EHRs), accessed via mobile devices or desktop computers. mHealth applications were primarily used for screening, clinical calculations, teaching, drug reference, reporting, and information retrieval. Tools that were mostly used in the US included mental health screening applications, surgical risk calculators, neonatal sepsis and bilirubin calculators, and MDCalc to support specific clinical tasks. In Tanzania, the Afya Supportive Supervision System (AfyaSS) was used for internal supervision, while the weekly Integrated Disease Surveillance and Response (IDSR) system was used to generate epidemiological reports. Providers in Tanzania also used reference tools such as the Antiretroviral Therapy (ART) app and Gestogram, whereas providers in both settings used resources such as Drugs.com and Medscape. To access current evidence, providers in the US used the United States Preventive Services Task Force (USPSTF) resources, and AI-based tools, including UpToDate, and OpenEvidence, and ChatGPT, while Tanzanian providers reported using ChatGPT more frequently.

### Perceived barriers and facilitators to use of FluidCalc

Using a CFIR-guided approach, we identified facilitators and barriers across relevant domains in both settings. Emergent themes were mapped to the specific CFIR domains and constructs, as shown in Table 3.

**Table 3.**
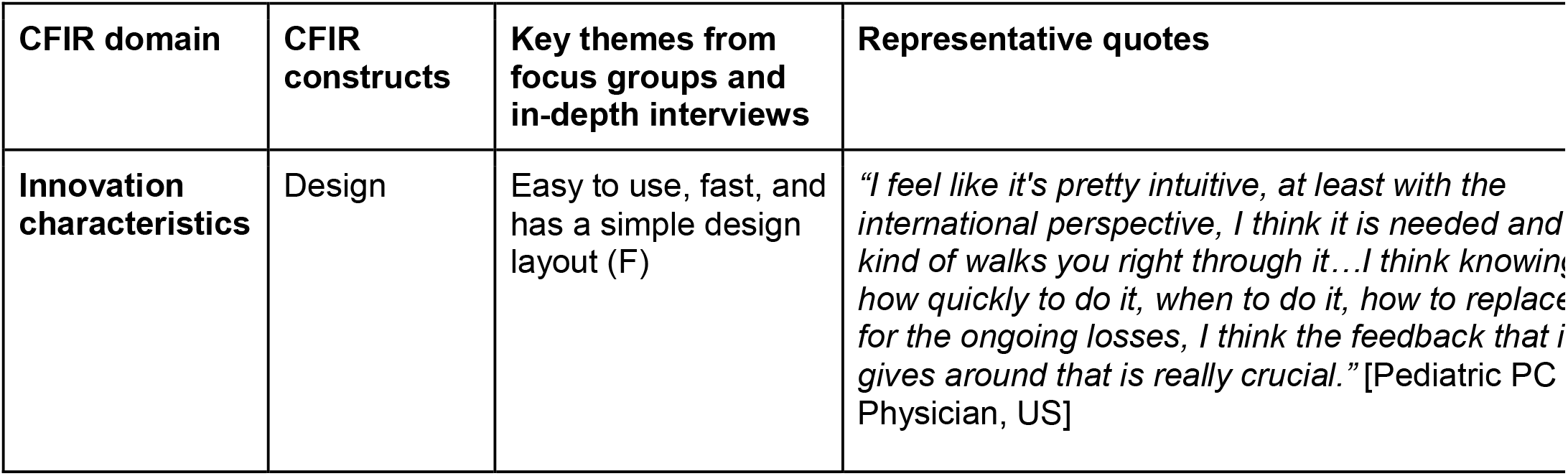
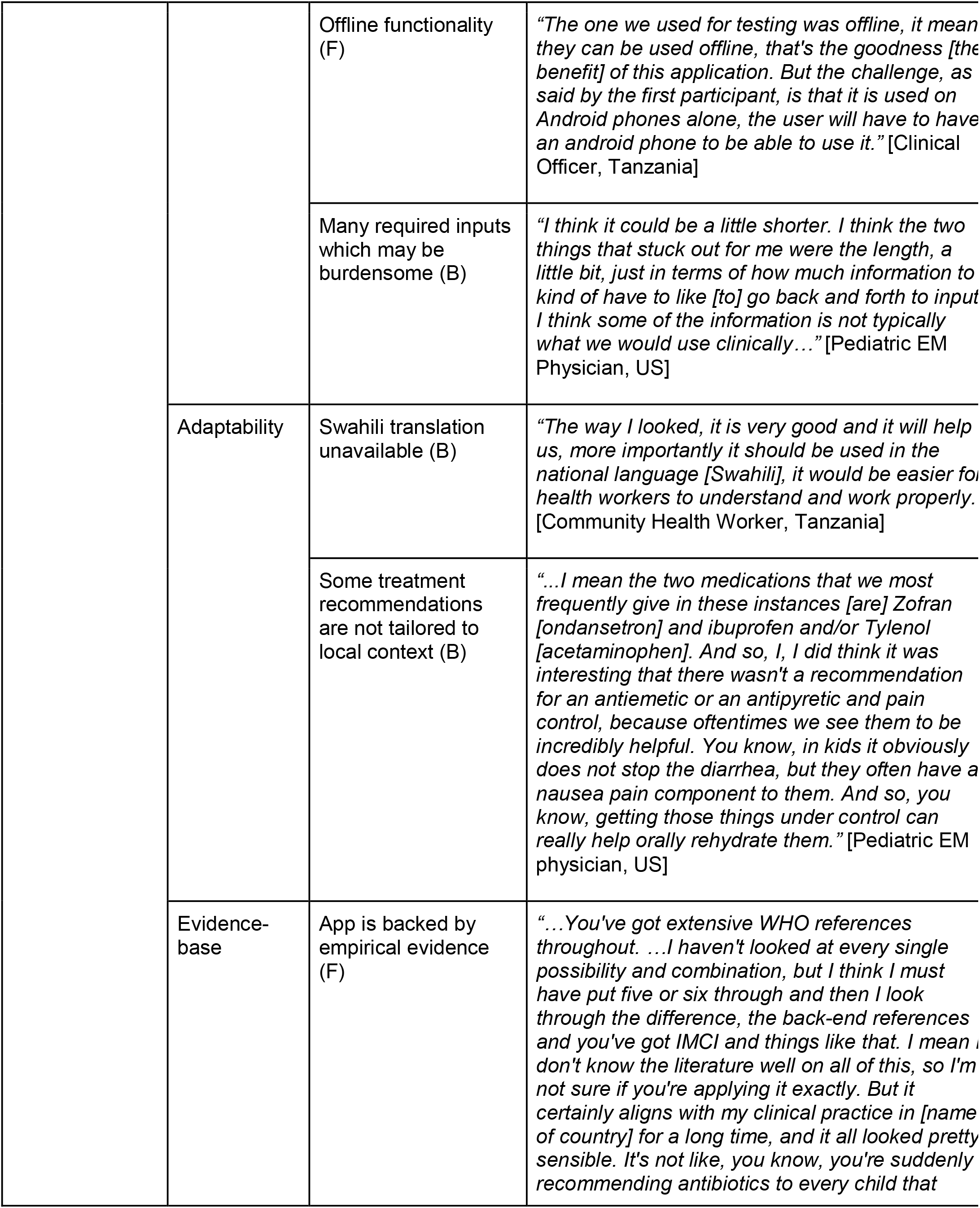
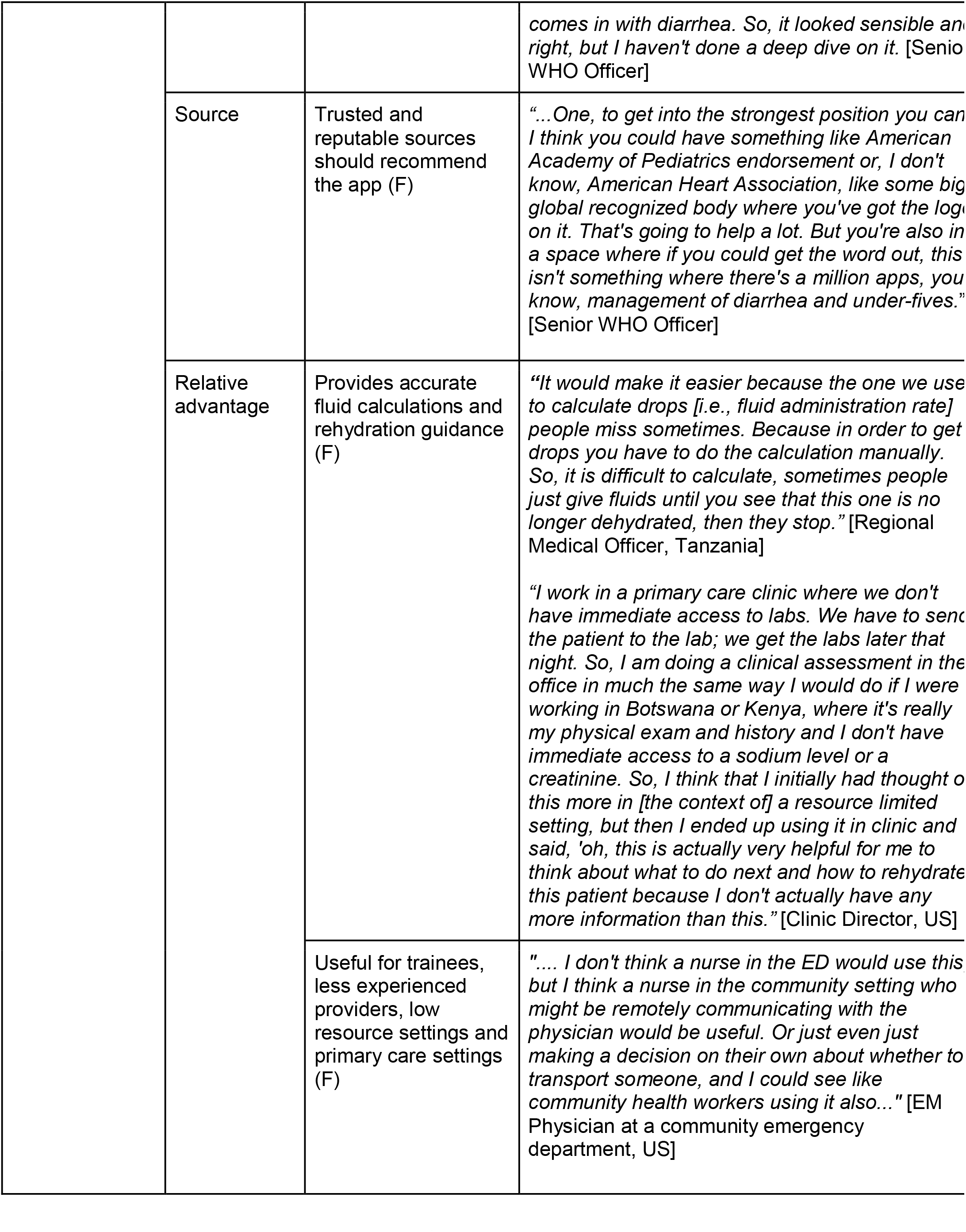
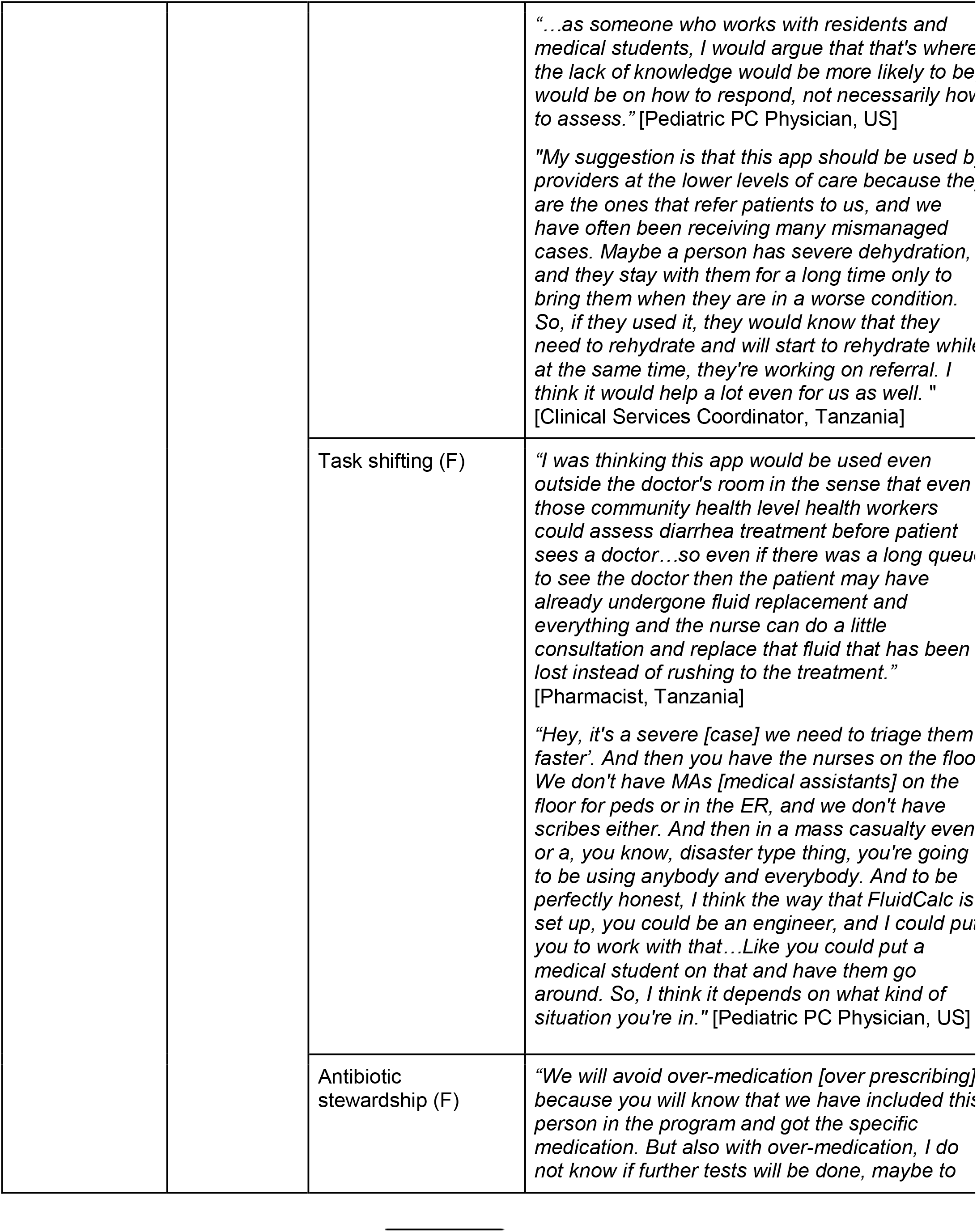
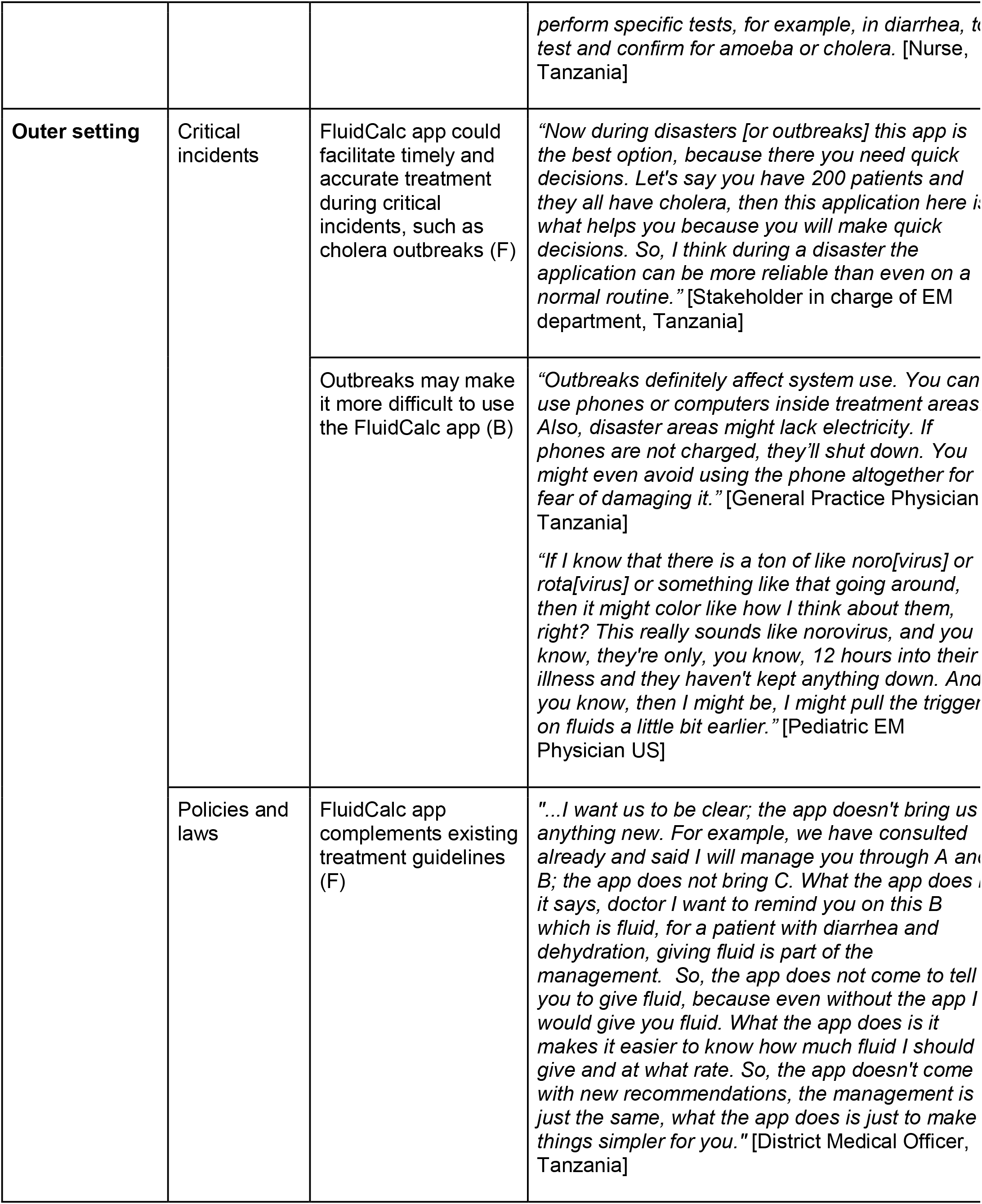
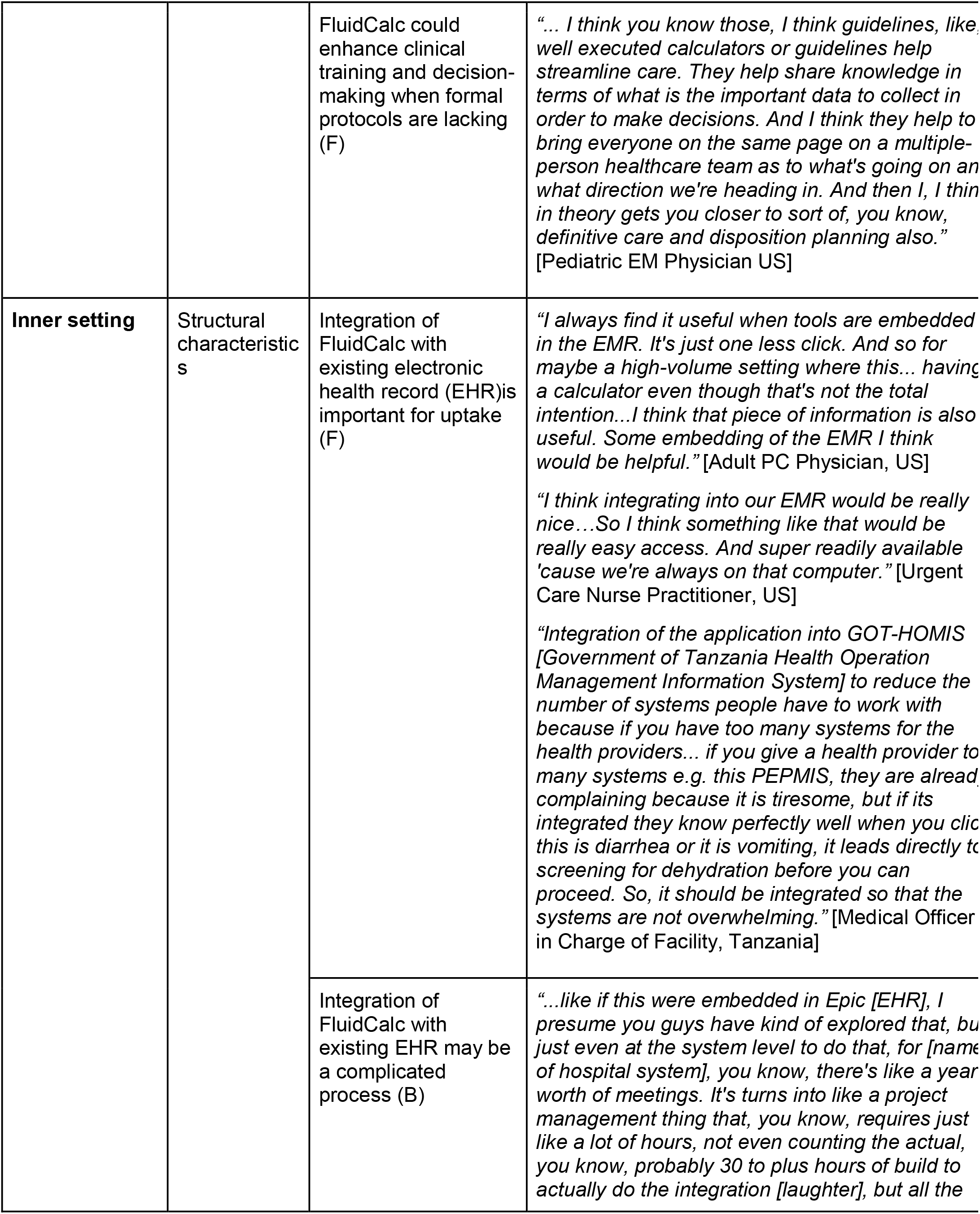
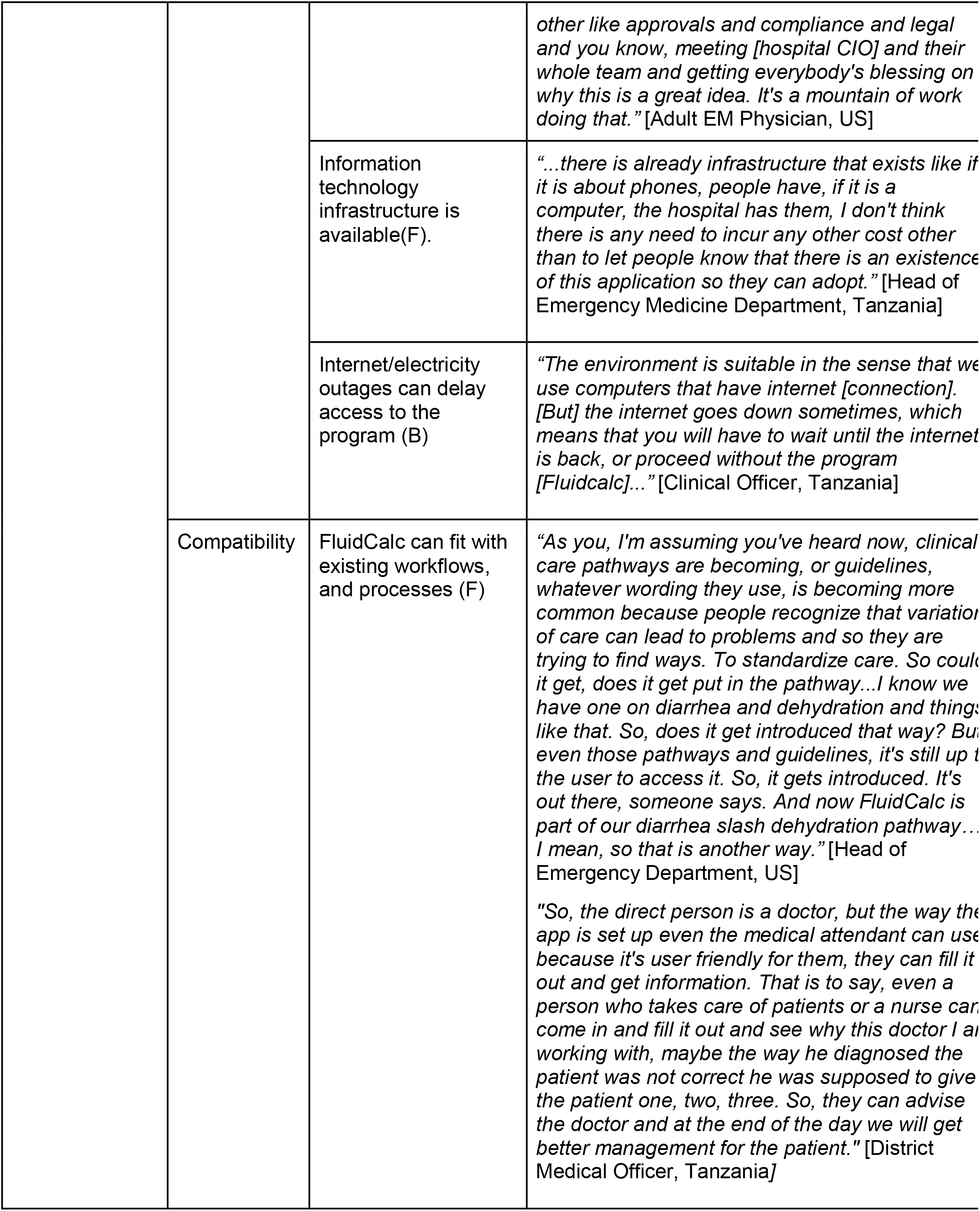
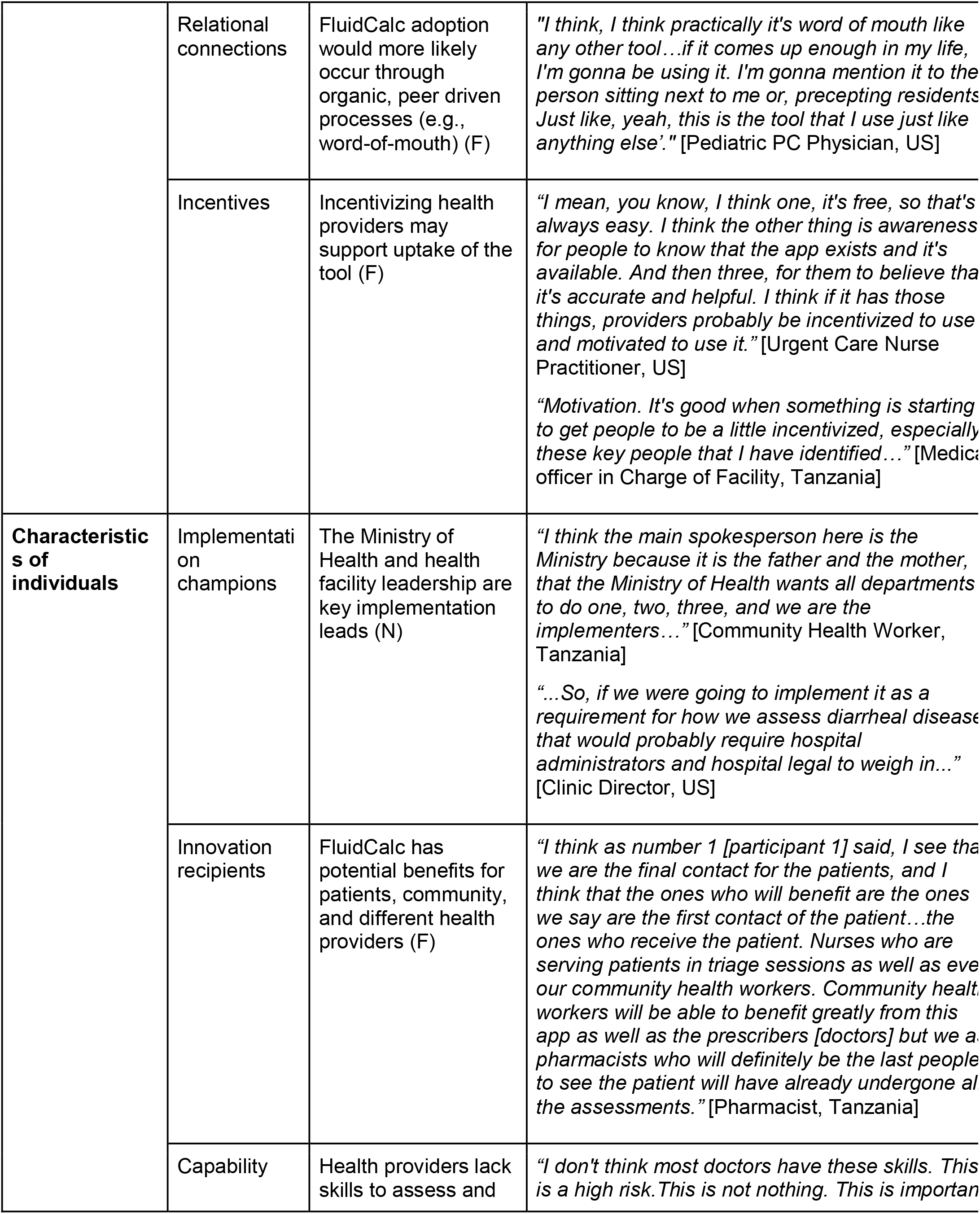
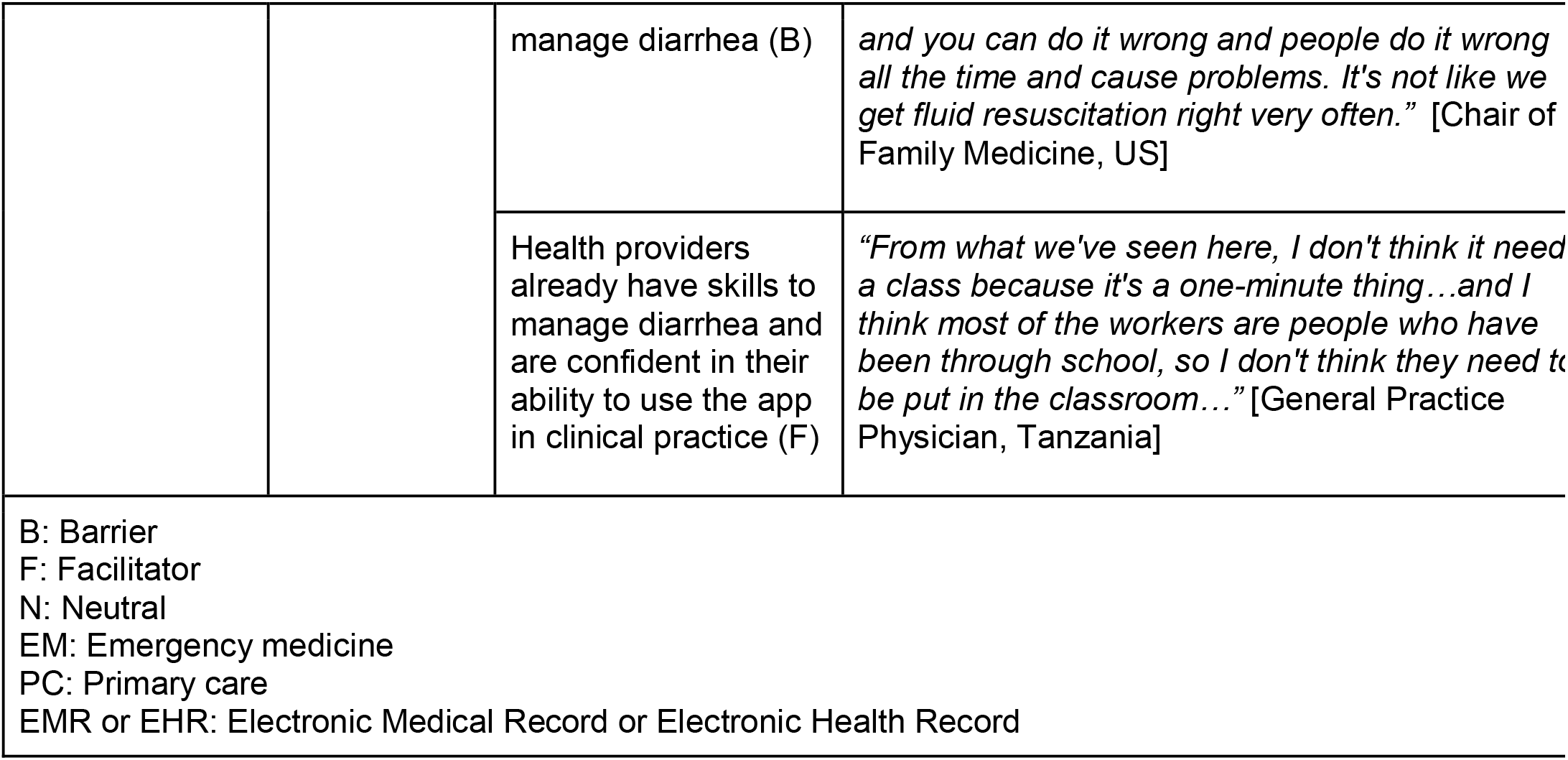
Summary of barriers and facilitators for implementation.

### Innovation characteristics

#### Facilitators

Participants from both settings reported that FluidCalc was easy to use, with a simple and intuitive interface. The app’s offline functionality was universally valued, and participants appreciated rapid functionality without delays or lags, making it efficient for use in busy clinical settings. Participants identified several advantages to using the app versus their current practice, including faster and more accurate fluid rehydration recommendations, support for rapid assessment and timely referrals, and facilitation of initiating guideline-adherent treatment at the community level. Participants from both settings also perceived the tool as valuable for pediatric care, supporting less experienced providers, serving as a teaching aid, and assisting with ongoing hydration management in inpatient and observation settings. Additionally, participants across all groups described FluidCalc as an important tool for supporting clinical decision-making and reducing reliance on less evidence-based approaches to diarrheal rehydration management. Healthcare providers across both settings agreed that FluidCalc would be particularly beneficial in low-resource settings, where accurate fluid management is critical for optimizing limited resources and supporting antibiotic stewardship.

Primary care providers in the US felt the app would be most useful for making disposition decisions (i.e. manage at home as outpatient versus refer for higher level of care) in patients with borderline or moderate dehydration, particularly in primary outpatient settings.

Tanzanian health providers reported that the treatment recommendations were consistent with national guidelines and their own clinical judgment. For instance, pharmacists appreciated the app’s precise fluid calculations and clear guidance on administering oral and intravenous fluids. Despite expressing confidence in the app’s recommendations, most providers, particularly physicians in both settings, emphasized that they would use FluidCalc to complement rather than replace their clinical judgment.

#### Barriers

Although participants generally considered the treatment recommendations to be evidence based, providers in both Tanzania and the United States reported that some recommendations did not reflect local clinical practice. US providers noted the absence of commonly used medications for acute gastroenteritis such as ondansetron (Zofran), acetaminophen and ibuprofen, unavailability of important clinical considerations such as vomiting and response to therapy, and uncertainty about the relevance of zinc supplementation in their setting. Providers in both countries also expressed concerns that the antibiotic recommendations did not sufficiently reflect local treatment guidelines or antimicrobial resistance patterns. They recommended expanding the antibiotic guidance while clearly emphasizing that antibiotics should only be prescribed when clinically indicated, informed by laboratory testing when available, and consistent with national treatment guidelines or facility- specific antibiotic protocols to promote appropriate antibiotic stewardship.

Health providers in Tanzania expressed concerns that the application did not adequately account for patient-specific factors, such as pregnancy and chronic comorbidities, including diabetes, cardiovascular disease, and hypertension, which could influence treatment decisions. Some participants were also uncertain about the rationale for including Vitamin A recommendations. Additional concerns related to implementation included difficulties downloading software updates in areas with unreliable internet connectivity, limited availability across mobile operating systems, and the absence of a Swahili version. A small number of participants also raised concerns about patient data security and the management of stored information. Some providers also noted that patients might misinterpret clinicians’ use of a mobile phone during consultations if the purpose of the application was not clearly explained.

In the US, participants identified several usability challenges, including the lack of a clear statement of the app’s purpose and intended users, the number of variables required for data entry, reliance on clinical variables that cannot be automatically populated from the electronic health record, a non-intuitive fluid rehydration interface, and lengthy additional information content that may be impractical to reference during busy clinical shifts. Many providers also felt that FluidCalc had limited utility for patients who were clearly well or critically ill and would be most valuable for intermediate-acuity cases. Experienced physicians noted that they would be less likely to rely on the app because they primarily use clinical judgment developed through experience.

### Outer setting

#### Facilitators

Participants consistently highlighted FluidCalc’s potential to improve care during outbreaks and other critical incidents by enabling timely fluid calculations and rapid clinical decision-making. Respondents believed the app could support task shifting by assisting less experienced providers, guiding triage, and facilitating clinical decision-making in high-stress or unfamiliar situations. Some providers also suggested that FluidCalc could support systematic data collection for resource allocation and could be adapted for telehealth consultations to facilitate remote assessment of large patient volumes or enable simplified patient self-triage based on symptom screening. Offline functionality was considered essential for use in field settings and during emergencies.

Participants in Tanzania generally agreed that existing policies and clinical protocols would not hinder implementation, instead they viewed FluidCalc as a tool that could complement existing guidelines by making them more accessible and reducing the need to consult printed treatment manuals during patient care. Respondents also emphasized that FluidCalc should be aligned with the Ministry of Health and World Health Organization guidelines to promote clinician confidence and facilitate adoption.

Participants in the US considered FluidCalc most applicable in primary care outpatient settings and during diarrhea outbreaks where rapid assessment and treatment decisions are required, rather than for patients who have already been stabilized in the emergency department or inpatient ward. They also viewed the app as a valuable resource for clinical training and decision support when formal protocols are unavailable. Additionally, respondents noted that app use would depend partly on personal workflow, with most indicating that they use tools on a second screen at their station or on their phone, with little barrier to using external tools on personal devices outside of the EHR system.

### Barriers

Some participants in Tanzania and the US felt that outbreaks or other critical incidents could actually make FluidCalc more difficult to use because of restrictions on bringing mobile phones into treatment areas, limited time for data entry, large patient volumes, lack of electricity and other infrastructure, and concerns about nosocomial infections. Specific to the US context, some participants indicated that changes in clinical management during outbreaks could reduce the need for FluidCalc. For instance, relying more on clinical gestalt and epidemiologic patterns (e.g., “what’s going around”) rather than formal tools, incorporating travel history and exposures earlier in the decision-making process, and shifts in care for more severe cases.

Additionally, US participants identified health facility workflows, clinician practice patterns, and patient preferences for oral rehydration strategies or the use of nasogastric (NG) versus intravenous (IV) fluids, as potential barriers to implementation because they influence how the app’s treatment recommendations could be applied in practice. For instance, emergency physicians caring for adult patients noted that nurses may find it difficult to administer rapid large volumes sometimes recommended by the app because they may not be feasible within existing clinical settings.

### Inner setting

#### Facilitators

Tanzanian health providers identified dehydration among patients with diarrhea as a common problem in Tanzania and emphasized the need for a tool such as FluidCalc in health facilities. FluidCalc could be integrated at multiple points within the diarrhea management workflow, with minimal interruption to the usual organization of tasks and responsibilities across different provider groups.

Health providers in both settings indicated that integration of FluidCalc CDST with existing EHR systems was a key facilitator of implementation. Integration of FluidCalc into GoT- HoMIS, Tanzania’s health information system, was identified as an important strategy to support adoption by reducing provider workload and preventing duplication of tasks, particularly in high- volume settings where health providers already use multiple digital tools. Similarly, participants in the US noted that integrating FluidCalc recommendations into existing electronic medical record workflows on Epic, including best practice advisory (BPA) alerts, could enhance usability and facilitate implementation, provided it is technically feasible.

### Barriers

Respondents identified several challenges related to integrating FluidCalc into existing digital systems. In Tanzania, integrating FluidCalc into GoT-HOMIS would require access to computers, such as laptops or desktop computers, whereas use as a standalone application would depend on the availability of smartphones or tablets, reliable internet connectivity, and sufficient data for app installation and software updates. Participants in the United States noted that integration into Epic EHR would require considerable time and multiple institutional approvals. Several respondents in the US also questioned whether importing data from the EHR into FluidCalc would save time or reduce clinician workload and raised concerns about the accuracy of existing EHR data, noting that documentation errors could affect the app’s recommendations.

Consequently, whereas some US health providers preferred optional, workflow-aligned integration within existing smart sets or clinical pathways rather than automated or mandatory use, some Tanzanian health providers favored mandatory implementation supported by regular inspections, supervision, and checklists to monitor use. US participants also noted that electronic medical record (EMR)-based alerts are frequently ignored, which could limit adoption if FluidCalc were implemented in this way.

### Characteristics of individuals

#### Facilitators

Most respondents expressed confidence in their ability to use FluidCalc in clinical practice. Participants in both settings reported that they had access to technology infrastructure including phones, tablets and computers at their workplace. Some health facilities in Tanzania also had generators that would support continued app use during power outages. Participants generally felt that their colleagues would be receptive to the app because it simplifies clinical decision-making.

In Tanzania, participants reported that the app may provide improved access to care and reduced illness severity. For health providers, FluidCalc could support task shifting, reduce the burden on physicians by enabling other healthcare workers, such as nurses, pharmacists, or community health workers, to assess dehydration and refer patients to hospital, and help providers remember and apply treatment guidelines. Participants also noted potential benefits for the government through improved patient tracking, optimized medication use, and reduced reliance on overburdened healthcare staff.

The providers identified as most likely to benefit from FluidCalc in the US included those caring for pediatric patients and healthcare providers in primary care settings and low-resource settings (such as rural, frontier, or tribal areas or critical access hospitals). Respondents also identified advantages for less experienced providers and trainees in making decisions regarding oral versus intravenous rehydration in particular. Community providers lacking pediatric expertise were also considered likely beneficiaries, especially in health facilities with lower pediatric patient volume. Some participants suggested that the app could also guide patients on when oral hydration at home may be appropriate to prevent unnecessary healthcare visits.

### Barriers

However, some providers in Tanzania noted that providers who do not routinely manage patients with diarrhea would require training to use the application. Some stakeholders in both settings also felt that additional training would be necessary to support effective use of the app. In US providers suggested that adoption would depend on reliable loading times and minimizing the workflow burden associated with entering multiple data points. They also recommended improvements such as the ability to leave some fields blank to enhance usability and support uptake.

### Implementation process

#### Facilitators

Participants in Tanzania reported that official endorsement of FluidCalc by facility leadership and support from the medical officer in charge and IT staff were key factors to increase clinician trust and uptake. Adoption is influenced by official recommendations or guidelines issued by the Ministry of Health, which are disseminated through regional and district administrative structures. Regional Referral Hospital Management Teams oversee approval and coordination at referral hospitals, while TAMISEMI and local government authorities review and approve implementation in district hospitals, health centers, and dispensaries. Regional leaders function as the enforcers of policies and guidelines approved by the MoH. Participants proposed several strategies to promote FluidCalc uptake, including establishing monthly targets for app use (e.g., the number of patients with some or severe dehydration managed using FluidCalc) to monitor implementation progress. They also recommended publicizing FluidCalc through continuing medical education (CME) and sensitization activities to increase awareness among health providers and communities.

US providers had similar views, they reported that the uptake of FluidCalc would depend on both individual and institutional factors. At the individual level, providers could choose to use the app independently with minimal barriers. However, broader integration, particularly through the EHR, would require institutional leadership and multiple levels of approval, including review by information technology (IT) teams, compliance, and quality improvement committees, physician informatics groups, interdisciplinary clinical review structures, and executive stakeholders.

Participants in the US also indicated that adoption would more likely occur through organic, peer-driven processes, e.g., word of mouth and use in teaching or clinical workflows. Some participants suggested that integration into commonly used platforms, e.g., UpToDate or MDCalc, could increase exposure and use (of note, the DHAKA and NIRUDAK models have since been added to MDCalc). Endorsements from reputable organizations such as the American Academy of Pediatrics were also thought to be helpful in facilitating uptake of the app, as health providers often place greater trust in recommendations and tools supported by well- established professional organizations. Successful implementation would also require demonstrating the tool’s clear purpose, utility and value. Respondents reported that this would involve identifying a strong use case such as pediatric patients or low-resource settings, intended users, demonstrating improvements in efficiency or reductions in disposition time and showing where FluidCalc outputs differ from clinician gestalt. Providing pre- and post-implementation data was also viewed as important for demonstrating impact and supporting uptake.

Additional strategies to support uptake in both settings included adding practical guidance, such as oral rehydration solution (ORS) preparation instructions, clarifying how inputs are used in calculations, and providing references to the empirical studies underlying the app’s algorithms and recommendations to reduce perceptions of the app as a “black box.” Participants also recommended expanding the app’s functionality by adding follow-up or stepped-care guidance for patients who do not improve and exploring telehealth or partially patient-facing versions of the tool.

## Discussion

Complementing prior implementation research conducted in Bangladesh our study demonstrates that FluidCalc CDST may have clinical utility in both high-resource and low-resource settings globally [25]. Implementation will require adaptation to local clinical workflows, treatment guidelines, common disease etiologies, and health system capacity. Providers reported that FluidCalc was easy to use and could support faster and more accurate management of patients with acute diarrhea. Specific feedback on application design and user experience will be reported separately.

Diarrhea management guidelines vary across and within healthcare settings. In Tanzania, the WHO IMAI and IMCI algorithms are commonly used for adults and children, respectively [30,31]. Tanzanian health providers reported reliance on paper-based WHO guidelines and manual fluid calculations, which they highlighted as often time-consuming and prone to error. Implementation of FluidCalc in Tanzania was positively viewed as a way to facilitate more rapid and accurate assessment of dehydration severity, improve adherence to evidence-based standards of care, and optimize resource utilization. The CDST allows users to toggle between the NIRUDAK model/DHAKA score and the WHO method for dehydration assessment, supporting use of more accurate prediction models while maintaining compatibility with current standards of care. Studies evaluating electronic WHO guidelines in low-resource settings have demonstrated improvements in dehydration assessment, more individualized fluid resuscitation, and reduced unnecessary intravenous fluid use [32,33].

Nearly all US health providers across all health facility types reported that there were no standardized clinical guidelines for diarrheal dehydration management. FluidCalc was identified as a valuable tool to address this gap. Health providers also indicated that FluidCalc may be greatly beneficial in primary care settings, serve as a teaching and decision-support resource for trainees and less-experienced clinicians, and for assessing patients with intermediate acuity where management decisions are less straightforward. The use of mHealth tools for clinical teaching is established in both high- and low-resource settings. For example, smartphone- based applications for just-in-time teaching by emergency medicine residents for medical students in the US have been shown to increase the frequency and quality of teaching, improve diagnostic documentation, and improve the accuracy of diagnostic coding. Resident physicians in resource-limited settings use smartphones with point-of-care tools to access medical information at the bedside and for self-directed learning [34–36].

Stakeholders and healthcare providers provided feedback on treatment recommendations, including fluid deficit calculations, fluid resuscitation, antibiotics, zinc, and vitamin A supplementation. Most participants considered the fluid deficit and resuscitation recommendations to be accurate and consistent with their clinical judgment. Across both settings, participants indicated that antibiotic recommendations should reflect local prescribing patterns. In Tanzania, where bacterial etiologies are more prevalent, providers recommended tiered antibiotic recommendations to promote appropriate use, accommodate patient preferences, and provide alternatives during drug shortages. In contrast, US providers perceived less value in the app’s antibiotic decision support because prescribing was already guided by established institutional protocols and most cases of acute diarrhea were attributed to viral pathogens. Additionally, international guidelines for use of zinc and vitamin A were not standard practice in diarrhea management due to the very low prevalence of these vitamin deficiencies in the US context. Previous studies have demonstrated that clinical decision support tools can improve adherence to guideline-recommended antibiotics and promote antibiotic stewardship [32,33,35,37]. Tailoring antibiotic recommendations to local epidemiology and prescribing practices may optimize antimicrobial treatment in Tanzania and other similar settings. Recommendations for vitamin A and zinc supplementation were considered relevant in Tanzania, where micronutrient deficiencies are more common among children. In contrast, US providers suggested including medications such as antiemetics and analgesics that are more routinely used in their practice.

While FluidCalc CDST is currently available on Android and iOS smartphones, web browsers, and MDCalc, integration into existing EHRs was identified as a strategy to improve uptake and use. Embedding the CDST within EHR systems could streamline clinical workflows by minimizing duplicate data entry and enabling automatic population of relevant patient information. EHR integration may also facilitate retention and retrieval of clinical data for disease reporting and surveillance, reduce data-entry burden and app fatigue among health providers, and allow providers without smartphones or those who prefer not to use phones during their shift to access FluidCalc from workplace computers. Participants noted that several CDSTs have already been embedded within Epic, including tools for lung cancer screening. However, embedding the FluidCalc CDST within EHR systems may be challenging, as customization is required to fit back-end system capabilities while preserving the integrity of underlying FluidCalc algorithms. Automatic population of patient information would also require interoperability between the CDST and EHR. Integration of the Kidney Failure Risk Equation and Statin Choice decision aid demonstrates the feasibility of incorporating CDSTs into complex EHR systems [38]. Further research is needed to identify effective approaches for integrating FluidCalc CDST into EHR systems across diverse healthcare settings.

Implementation requires not only technical adaptation but also context-specific considerations regarding clinical workflows, acceptability by providers, regulatory approvals, and clinician training and infrastructure support. Physicians in both settings expressed greater agency in adopting CDSTs and were less likely to require institutional approval. Other provider groups emphasized the need for formal approval from facility leadership or the Ministry of Health. Local champions and institutional leadership were identified as important for clinician buy-in. Physicians also preferred to retain autonomy over app use and described it as supportive rather than directive in clinical decision-making. These findings are consistent with our previous study in Bangladesh, which demonstrated that physicians and clinicians with existing confidence in managing diarrhea, intended to use the tool as an adjunct to clinical judgment and advocated for more directive guidance if used by non-physician providers [25]. Transparency regarding the evidence supporting FluidCalc algorithms and treatment recommendations, as well as patient data storage and use, was also considered essential for building trust in the tool.

Critical incidents, such as cholera or norovirus outbreaks, were identified as potential use cases for CDST. FluidCalc may support triage, facilitate rapid clinical decision-making in high-volume settings, and enable task shifting to nurses, community health workers, and other frontline providers. However, use during critical incidents may be limited by lack of electricity, limited access to electronic devices or internet connectivity, and challenges integrating digital tools into rapidly evolving clinical environments. In low resource settings with clinician shortages, CHWs could use the app to assess patients with diarrhea, identify those requiring referral for physician evaluation or facility-based fluid resuscitation, thus reducing physician workload. Previous studies have shown that CHWs can address health workforce shortages and, when adequately trained and supported, improve population health outcomes by linking patients in their community to care [25, 39].

### Limitations

Participants’ perceptions of the tool’s usability were based primarily on app demonstrations and limited individual use prior to interviews and focus groups. Some participants had not used the tool or experienced difficulties accessing it online. These findings therefore reflect initial exposure rather than extended use in clinical settings. The extent of adoption and utilization in practice remains to be determined. Fewer participants were recruited in some provider categories due to limited clinician availability. In Tanzania, all intended provider groups were represented. In the United States, a focus group with mid-level providers could not be conducted due to insufficient participant availability, potentially underrepresenting this group. Inclusion of a diverse range of stakeholders, from hospital leaders to international policymakers, strengthens the study by capturing implementation considerations across multiple levels of the health system.

## Conclusion

FluidCalc has the potential to improve diarrhea management in both low- and high- resource settings and can be implemented either as a standalone mHealth app or as an integrated component within an EHR system. Although several implementation barriers were identified, these do not outweigh the potential advantages of the FluidCalc clinical decision support tool. Rather, they highlight the need to tailor implementation strategies to local clinical workflows, and health system infrastructure. The implementation determinants identified in this study provide a foundation for future implementation across similar healthcare settings. Future research should evaluate implementation in routine clinical practice, including impact on clinical decision-making, adherence to antibiotic prescribing patterns, and specific patient outcomes.

## Data Availability

The dataset (which includes individual transcripts) is not publicly available due to confidentiality policies.

## List of abbreviations

AI: Artificial Intelligence
ART: Antiretroviral Therapy
BPA: Best Practice Advisory
CDSTs: Clinical Decision Support Tools
CFIR: Consolidated Framework for Implementation Research
CHW: Community Health Worker
DHAKA: Dehydration Assessing Kids Accurately
EHR: Electronic Health Record
EM: Emergency Medicine
EMR: Electronic Medical Record
FGD: Focus Group Discussion
IDI: In-depth Interview
IDSR: Integrated Disease Surveillance and Response
IMAI: Integrated Management of Adolescent and Adult Illness
IMCI: Integrated Management of Childhood Illness
IT: Information Technology
IV: Intravenous
LMIC: Low- and Middle-Income Countries
mHealth: Mobile Health
NG: Nasogastric
NIRUDAK: Novel Innovative Research for Understanding Dehydration in Adults and Kids
PC: Primary Care
USPSTF: United States Preventive Services Task Force
WHO: World Health Organization

## Declarations

### Ethics of approval and consent to participate

This study was approved by Muhimbili University of Health and Allied Sciences Research Review Committee/Ethical Review Committee Protocol Approval: MUHAS-REC-02-2025-2619, Brown University Institutional Review Board Protocol Approval #: STUDY00000598 & STUDY00000977, in accordance with the Declaration of Helsinki.

### Competing interests

The authors declare that they have no competing interests.

### Funding

Funding was provided through grants from the National Institute for Health (NIH) National Institute for Diabetes and Diarrheal and Kidney Diseases (NIDDK), (PI Levine, R01DK116163).

### Authors contributions

ACL contributed to conceptualization, overall project supervision and coordination, and acquired grant funds from the main sponsor. ACL and KPM provided project supervision at individual study sites, and JC provided project administration. ACL, SCG, RKR, CD and KPM designed the research study. JC, SCG, RKR, RL and ACL designed the protocol. JC, RKR, SCG, RL, MS, RR, FD, KPM and ACL contributed to data collection. JC, RL, MS, RR, FD, DY and VS contributed to data cleaning. JC, RKR, SCG, RL, DY, VS and ACL conducted the data analysis. JC wrote the original draft of the manuscript. All authors reviewed and provided feedback on the draft and approved the final draft of the manuscript. All authors had full access to all the data in the study and had final responsibility for the decision to submit for publication.

## Acknowledgements

The authors thank all study participants and the study staff at Muhimbili University for Health and Allied Sciences, Brown University and Brown University Health who were instrumental in collecting the data used in this study.

## Notes

### Competing Interest Statement

The authors have declared no competing interest.

### Author Declarations

Ethics committee/IRB of Muhimbili University of Health and Allied Sciences and Brown University gave ethical approval for this work

## References

1. Wang T, Wang L, Sang X, Ren Y, Xu T, Huang Q, Xiao A, Lu W, Li H, Li S, Wu X. Regional and age-specific global trends associated with infectious diarrhea in children under 14 years old caused by pathogenic microorganisms in 2021. Front Med. 2025; 12:1676249. doi:10.3389/fmed.2025.1676249.

2. Liang D, Wang L, Liu S, Li S, Zhou X, Xiao Y, et al. Global incidence of diarrheal diseases— an update using an interpretable predictive model based on XGBoost and SHAP: a systematic analysis. Nutrients. 2024;16(18):3217. doi:10.3390/nu16183217.

3. Zhao W-Z, Wang J-Y, Zhang M-N, Wu S-N, Dai W-J, Yang X-Z, et al. Global burden of diarrhea disease in the older adult and its attributable risk factors from 1990 to 2021: a comprehensive analysis from the Global Burden of Disease Study 2021. Front Public Health. 2025; 13:1541492. doi:10.3389/fpubh.2025.1541492.

4. Meisenheimer ES, Epstein C, Thiel D. Acute diarrhea in adults. American Family Physician. 2022;106(1):72–80.

5. Munos MK, Fischer Walker CL, Black RE. The effect of oral rehydration solution and recommended home fluids on diarrhoea mortality. Int J Epidemiol. 2010;39(Suppl 1): i75– i87. doi:10.1093/ije/dyq025.

6. Hartling L, Bellemare S, Wiebe N, Russell KF, Klassen TP, Craig WR. Oral versus intravenous rehydration for treating dehydration due to gastroenteritis in children. Cochrane Database Syst Rev. 2006;(3):CD004390. doi: 10.1002/14651858.CD004390.pub2.

7. Levine AC, Barry MA, Gainey M, Nasrin S, Qu K, Schmid CH, et al. Derivation of the first clinical diagnostic models for dehydration severity in patients over five years with acute diarrhea. PLoS Negl Trop Dis. 2021;15(3): e0009266. doi: 10.1371/journal.pntd.0009266.

8. Daley SF, Avva U. Pediatric dehydration. In: StatPearls. Treasure Island (FL): StatPearls Publishing; 2026.

9. Tsegaye AT, Pavlinac PB, Walson JL, Tickell KD. The diagnosis and management of dehydration in children with wasting or nutritional edema: a systematic review. PLOS Glob Public Health. 2023;3(11): e0002520. doi: 10.1371/journal.pgph.0002520.

10. Gainey M, Barry M, Levine AC, Nasrin S. Developing a novel mobile health (mHealth) tool to improve dehydration assessment and management in patients with acute diarrhea in resource-limited settings. Rhode Island medical journal (2013). 2019 Sep 3;102(7):36.

11. Pringle K, Shah SP, Umulisa I, Mark Munyaneza RB, Dushimiyimana JM, Stegmann K, Musavuli J, Ngabitsinze P, Stulac S, Levine AC. Comparing the accuracy of the three popular clinical dehydration scales in children with diarrhea. International journal of emergency medicine. 2011 Sep 9;4(1):58. doi:10.1186/1865-1380-4-58

12. Falszewska A, Dziechciarz P, Szajewska H. Diagnostic accuracy of clinical dehydration scales in children. European Journal of Pediatrics. 2017 Aug;176(8):1021–6. doi:10.1007/s00431-017-2942-8.

13. Levine AC, Glavis-Bloom J, Modi P, Nasrin S, Rege S, Chu C, Schmid CH, Alam NH. Empirically derived dehydration scoring and decision tree models for children with diarrhea: assessment and internal validation in a prospective cohort study in Dhaka, Bangladesh. Global Health: Science and Practice. 2015 Sep 10;3(3):405–18. doi:10.9745/GHSP-D-15-00097.

14. Barry MA, Qu K, Gainey M, Schmid CH, Garbern SC, Nasrin S, Alam NH, Lee JA, Nelson EJ, Rosen R, Levine AC. Derivation and internal validation of a score to predict dehydration severity in patients over 5 years with acute diarrhea. The American Journal of Tropical Medicine and Hygiene. 2021 Aug 16;105(5):1368. doi:10.4269/ajtmh.21-0143

15. Agarwal S, Glenton C, Tamrat T, Henschke N, Maayan N, Fønhus MS, Mehl GL, Lewin S. Decision-support tools via mobile devices to improve quality of care in primary healthcare settings. Cochrane Database of Systematic Reviews. 2021(7). doi: 10.1002/14651858.CD012944.pub2.

16. White A, Thomas DS, Ezeanochie N, Bull S. Health worker mHealth utilization: a systematic review. CIN: Computers, Informatics, Nursing. 2016 May 1;34(5):206–13. doi:10.1097/CIN.0000000000000231.

17. Amoakoh HB, Klipstein-Grobusch K, Grobbee DE, Amoakoh-Coleman M, Oduro-Mensah E, Sarpong C, et al. Using mobile health to support clinical decision-making to improve maternal and neonatal health outcomes in Ghana: insights of frontline health worker information needs. JMIR Mhealth Uhealth. 2019;7(5): e12879. doi:10.2196/12879.

18. Amoakoh-Coleman M, Borgstein AB, Sondaal SF, Grobbee DE, Miltenburg AS, Verwijs M, et al. Effectiveness of mHealth interventions targeting health care workers to improve pregnancy outcomes in low- and middle-income countries: a systematic review. J Med Internet Res. 2016;18(8): e226. doi:10.2196/jmir.5533.

19. Bilal S, Nelson E, Meisner L, Alam M, Al Amin S, Ashenafi Y, et al. Evaluation of standard and mobile health-supported clinical diagnostic tools for assessing dehydration in patients with diarrhea in rural Bangladesh. Am J Trop Med Hyg. 2018;99(1):171. doi:10.4269/ajtmh.17-0648

20. McLaughlin M, Metiboba L, Giwa A, Femi-Ojo O, Ravi N, Mahmoud NM, et al. Adherence to integrated management of childhood illness (IMCI) guidelines by community health workers in Kano State, Nigeria through use of a clinical decision support (CDS) platform. BMC Health Serv Res. 2024;24(1):953. doi:10.1186/s12913-024-11245-z.

21. Braun R, Catalani C, Wimbush J, Israelski D. Community health workers and mobile technology: a systematic review of the literature. PLoS One. 2013;8(6): e65772. doi: 10.1371/journal.pone.0065772

22. Feroz A, Jabeen R, Saleem S. Using mobile phones to improve community health workers performance in low-and-middle-income countries. BMC public health. 2020 Jan 13;20(1):49. doi:10.1186/s12889-020-8173-3

23. Mahmood H, McKinstry B, Luz S, Fairhurst K, Nasim S, Hazir T, et al. Community health worker-based mobile health (mHealth) approaches for improving management and caregiver knowledge of common childhood infections: a systematic review. J Glob Health. 2020;10(2):020438. doi:10.7189/jogh.10.020438.

24. Addotey-Delove M, Scott RE, Mars M. Healthcare workers’ perspectives of mHealth adoption factors in the developing world: scoping review. Int J Environ Res Public Health. 2023;20(2): 1244.R doi:10.3390/ijerph20021244.

25. Rosen RK, Garbern SC, Gainey M, Lantini R, Nasrin S, Nelson EJ, et al. Designing a novel clinician decision support tool for the management of acute diarrhea in Bangladesh: formative qualitative study. JMIR Hum Factors. 2022;9(1): e33325. doi:10.2196/33325.

26. Levine AC, Gainey M, Qu K, Nasrin S, Sharif MB, Noor SS, et al. A comparison of the NIRUDAK models and WHO algorithm for dehydration assessment in older children and adults with acute diarrhoea: a prospective, observational study. Lancet Glob Health. 2023;11(11): e1725–e1733. doi:10.1016/S2214-109X(23)00403-5.

27. Levine AC, Glavis-Bloom J, Modi P, Nasrin S, Atika B, Rege S, et al. External validation of the DHAKA score and comparison with the current IMCI algorithm for the assessment of dehydration in children with diarrhoea: a prospective cohort study. Lancet Glob Health. 2016;4(10): e744–e751. doi:10.1016/S2214-109X(16)30150-4.

28. Zakerabasali S, Ayyoubzadeh SM, Baniasadi T, Yazdani A, Abhari S. Mobile health technology and healthcare providers: systemic barriers to adoption. Healthc Inform Res. 2021;27(4):267–278. doi:10.4258/hir.2021.27.4.267

29. Rosen RK, Gainey M, Nasrin S, Garbern SC, Lantini R, Elshabassi N, et al. Use of framework matrix and thematic coding methods in qualitative analysis for mHealth: the FluidCalc app. Int J Qual Methods. 2023; 22:16094069231184123.

30. World Health Organization. IMAI district clinician manual: hospital care for adolescents and adults. 2nd ed. Geneva: World Health Organization; 2016. Available from: https://iris.who.int/bitstream/handle/10665/77751/9789241548281_Vol1_eng.pdf

31. World Health Organization. The treatment of diarrhea: a manual for physicians and other senior health workers. Geneva: World Health Organization; 2005. Available from: https://apps.who.int/iris/bitstream/10665/43209/1/9241593180.pdf

32. Khan AI, Mack JA, Salimuzzaman M, Zion MI, Sujon H, Ball RL, et al. Electronic decision support and diarrhoeal disease guideline adherence (mHDM): a cluster randomised controlled trial. Lancet Digit Health. 2020;2(5): e250–e258. doi:10.1016/S2589-7500(20)30062-5. doi:10.1016/S2589-7500(20)30062-5

33. Haque F, Ball RL, Khatun S, Ahmed M, Kache S, Chisti MJ, et al. Evaluation of a smartphone decision-support tool for diarrheal disease management in a resource-limited setting. PLoS Negl Trop Dis. 2017;11(1): e0005290. doi:10.1371/journal.pntd.0005290.

34. Ginsburg J, Sande M, Ahmed A, Moak J, Sudhir A, Mutter MK. A smartphone resource for just-in-time medical student teaching by emergency medicine residents. Cureus. 2024;16(9): e70117. doi:10.7759/cureus.70117.

35. Eudaley ST, Mihm AE, Higdon R, Jeter J, Chamberlin SM. Development and implementation of a clinical decision support tool for treatment of uncomplicated urinary tract infections in a family medicine resident clinic. J Am Pharm Assoc (2003). 2019;59(4):579– 585.

36. Chang AY, Ghose S, Littman-Quinn R, Anolik RB, Kyer A, Mazhani L, et al. Use of mobile learning by resident physicians in Botswana. Telemed J E Health. 2012;18(1):11–13. doi:10.1089/tmj.2011.0050. doi:10.1089/tmj.2011.0050

37. Abraham A, Shah R, Finette B, Sang EN, McLaughlin M, Mount-Finette E, et al. Enhancing quality of health care delivered to under 5 children in Kibra informal settlement of Nairobi, Kenya, by using mHealth platform with electronic clinical decision support system. Oxford Open Digit Health. 2025;3: oqaf030. doi:10.1093/oodh/oqaf030.

38. Alexiuk M, Elgubtan H, Tangri N. Clinical decision support tools in the electronic medical record. Kidney Int Rep. 2024;9(1):29–38.

39. Perry HB, Zulliger R, Rogers MM. Community health workers in low-, middle-, and high-income countries: an overview of their history, recent evolution, and current effectiveness. Annu Rev Public Health. 2014; 35:399–421.

